# The Effects of Pericapsular nerve group (PENG) block on Postoperative Recovery in Elderly Patients with Hip fracture: a study protocol for randomized, parallel controlled, double-blind trial

**DOI:** 10.1101/2021.03.14.21253545

**Authors:** Wei Luo, Quehua Luo, Jieting Wu, Jianhui Liang, Huiyi Wu, Yanhua Ou, Yuhui Li, Wuhua Ma

## Abstract

**Introduction:** Hip fracture is a common and serious emergency in the elderly, it is associated with severe pain, significant postoperative morbidity and mortality. Featuring peripheral nerve block in Enhanced recovery after surgery (ERAS) pathway may have significantly effect on shortening length of hospital stay, decreasing complications and costs, particular in improvement in dynamic pain and reducing the use of opioid. Pericapsular Nerve Group Block(PENG), suggested by Arango *et al*, may provide a effective blockade to the articular branches of the anterior hip joint,which innervate the most section of the hip capsule richly, with a potential motor-sparing effect.The purpose of this trail is to investigate whether PENG is effective to enhanced recovery in elderly patients with hip fracture.

**Methods and analysis:** This study will be a single centre,randomized, parallel controlled, double-blind trail. 92 elderly patients scheduled for hip fracture surgery will be divided into two groups randomly to receive ultrasound-guided PENG block or ultrasound-guided femoral nerve(FN) block. The primary outcome will be compare the quality of recovery-15(QoR-15) score at 24h postoperatively between two groups.The secondary outcomes include the strength of quadriceps, the visual analogue scale(VAS) at rest and on movement, the total morphine consumption, the rescue analgesic, the first time of postoperative out-of-bed mobilization, the complications.

**Ethics and dissemination:** This study has been approved by the Medical Science Research Ethics Committees of The First Affiliated Hospital of Guangzhou University of Chinese Medicine on 15 December 2020 (Reference K2020-110). The results of this study will be published in peer-reviewed international journals.

**Trial registration number:** ChiCTR2100042341.

## INTRODUCTION

The number of hip fractures in elder people is increasing steadily. In China, a study calculates, the annual incidence was 99.15 men and 177.13 women per 100,000 in 2016.^1^ And in United States, the rates of age-adjusted hip fracture for 2015 were higher than projected, resulting in an estimated increase of more than 11,000.^2^ The highest overall incidence is between 71 and 85 years old.^3^

The pain of hip fracture is serious, and VAS is more than 4 of 10 points,^4^ which is moderate to severe pain. It is seriously increase length of hospital stay and postoperative mortality.^5^ Early surgery is the definite treatment in most patients. It could control the pain, quicken the rehabilitation and decrease complications potentially.^6^ However the elderly often suffer from compromised organ reserve capacity, poor perioperative health status and varying systemic diseases. So, with the high postoperative morbidity and mortality,^7^ the safety of aging patients in perioperative period is full of challenges.

ERAS is a multimodal, multidisciplinary approach to the surgical patient in the perioperative period. There is good and increasing evidence that implementation of ERAS pathways can shorten length of hospital stay, decrease complications and costs for the patients undergoing hip surgery.^8-10^ And this multimodal pathway featuring peripheral nerve block may have significantly effect on fewer complications and perioperative outcomes after major orthopedic surgery. Comparing with tradional intravenous opioids during the initial postoperative period, peripheral nerve block can improve the dynamic pain control, quicken rehabilitation and reduce the use of opioids and the related adverse effects.^11-13^

With the development of the ultrasound guidance,peripheral nerve block is used frequently nowadays. Ultrasound guided femoral nerve(FN) block, fascia iliac block (FIB)and 3-in-1 FN block are popular peripheral nerve block techniques for hip fracture. However, as the obturator nerve(ON) and the accessory obturator nerve(AON) have often failed to exhibit adequate blockade from these blocks,the effect size is only moderate with decrease in the strength of quadriceps muscle.^14-16^

In order to improve the analgesia effect, according to the recent anatomical study by Short *et al*.^17^about the identification of relevant landmarks to the innervation of the anterior hip capsule,Arango *et al*.^18^ created a new regional technique for hip fractures: ultrasound-guided pericapsular nerve group (PENG)block. They concluded this technique could provide a effective blockade to the article branch of FN, ON and AON with a potential motor-sparing effect.

However, the case series of PENG block is small. And, without appropriate control groups, the outcomes have been limited. It is still unclear that whether PENG block is effect on the pain relief and rehabilitation in elderly patients with hip fractures within a standard care program. Therefore, a randomized, parallel controlled, double-blind trial will be set up, comparing the QoR-15 score at 24h postoperatively between two groups, to investigate whether PENG is effective to the enhanced recovery in elderly patients with hip fracture.

## METHODS AND ANALYSIS

### Primary aims

The primary aim of this trail is to investigate whether PENG is effective to the enhanced recovery in elderly patients with hip fracture.

### Secondary aims

The secondary aims are to investigate if PENG results in a motor-sparing effect,a reduction in the pain score at rest and on movement,a decrease in total morphine consumption and the rescue analgesic,an improvement in the first time of postoperative out-of-bed mobilization,and a decrease in complications.

### Trail design

This is a pragmatic, parallel group,randomized controlled, double-blind trial.The study will be carried out at The First Affiliated Hospital of Guangzhou University of Chinese Medicine and has been registered with the Chinese Clinical Trial Registry (ChiCTR2100042341). The flow diagram for this trial is presented in Figure 1.

**Figure 1.**
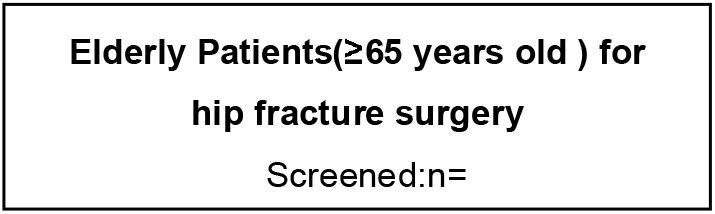
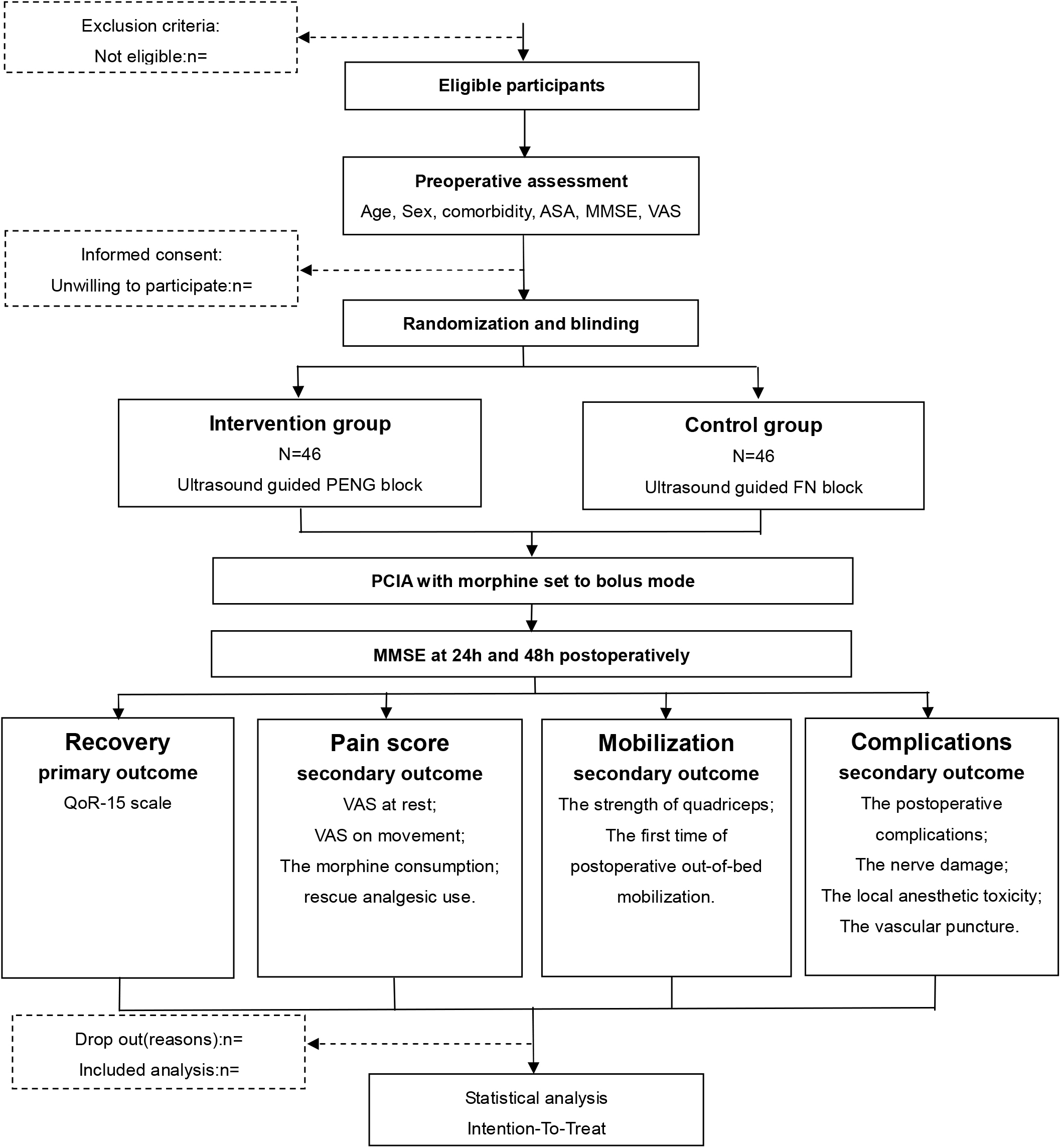
The flow diagram for this trial. ASA: American Society of Anesthesiologists class; MMSE: Mini-Mental State Examination; VAS: Visual analogue scale; PENG: Pericapsular nerve group; FN: Femoral nerve; PCIA: Patient controlled intravenous analgesia; QoR-15: quality of recovery-15.

### Eligibility criteria

#### Recruitment

Elderly patients(aged over 65 years old) with hip fracture who are selected for the hip surgery at The First Affiliated Hospital of Guangzhou University of Chinese Medicine will be recruited for this study.

#### Inclusion criteria

Patients will be included if they are cognitively intact, the American Society of Anesthesiologists physical status (ASA) I-□ and the body mass index(BMI) 18-30kg/m2.

#### Exclusion criteria

Patients will be excluded if they refuse to participate in the study, are allergies to analgesic drugs used in this study, dementia, multipal trauma, severe deafness and vision problems, communication difficulties, have an infection near the block site, are obesity (BMI>30kg/m2), have a significant clinically neurological,cardiovascular, renal and hepatic diseases(ASA □-□).

### Informed consent

According the inclusion criteria and the exclusion criteria,patients will be assessed strictly by the dedicated researchers the day before the surgery. Once patients is eligible, the researchers will give the full explanations to the eligible patients and their families about this study (including the implications, the known adverse effects, any risks participated in taking part and constraints of the protocol), and answer the questions about this study which the participants are interested in, to make sure they are clear about the significance of this study and eliminate their doubts. If patients agree to enrol, the informed consent will be obtained. The participants and their proxies will be asked to give written informed consent and sign personally. Of course, the participants can choose to withdraw from the trial freely without any reasons, and we will ask the agreement about the use of their data.

### Preoperative management

In accordance with the national guideline,^6^ patients in both groups will be received a standard care to accelerate recovery. The pathway includes rapid assessment from the Emergency Department, adequate pain control, assessment of bone health and falls, multidisciplinary management, surgical procedures and mobilization strategies. The ASA status classification will be evaluated and preoperative cognitive assessment will be performed by using the Mini-Mental State Examination (MMSE).^19^ The opioid will be avoided to use except for severe pain. Patients who participate in this study will avoid the use of premedication.

### Intraoperative management

After entering the operating room, continuous electrocardiogram, noninvasive intermittent blood pressure and pulse oxygen saturation will be monitored. After an intravenous is performed with an 18-gauge vein needle, lactated ringer solution will be dripped. The patients will be given 4 L / min of oxygen through the mask. In order to reduce the pain and fear during the nerve blockade, 0.5 ug/ kg of fentanyl will be given intravenously. The blockade of Both groups will be performed by the same experienced anesthesiologist. Before the local anesthesia, the anesthesiologist will open the envelops to get the sequence and perform the blockade according the allocation information. The anesthesia for the surgery will be performed after the block and managed by another anesthesiologists.In accordance with the assessment of patients, the anesthetic mode will be at the discretion of the anesthesiologist.

### Ultrasound guided pericapular nerve group block

With the participants are in a supine position, PENG block is performing as described by Arango *et al*.^18^ The anterior inferior iliac spine, the femoral artery(FA), the pectineus muscle, the iliopubic eminence, the iliopsoas muscle and tendon could be observed using a curvilinear low-frequency ultrasound probe. The puncture site will be set 0.5-1.0 cm away from the lateral of the ultrasound probe. After 2 ml of 1% lidocaine is injected for the local anesthesia, a 22-gauge, 80-mm needle will be inserted carefully in an in-plane approach from lateral to inner. Following the tip of the needle reaches the musculofascial between the tendon of the psoas muscle anteriorly and the pubic ramus posteriorly, a total volume 20mL of 0.375% ropivacaine will be slowly injected after negative aspiration every 5-mL.The ultrasound view of the fluid spread in the plane will be observed to ensure that the ropivacaine is injected right in the targeted location.

### Ultrasound guided femoral nerve block

With the patients are in a supine position, FN block is performed as described by Marhofer *et al*.^20^ A linear high-frequency probe will be used to visualize the FA and the FN. The puncture site will be set 0.5-1.0 cm away from the lateral of the ultrasound probe. After 2 ml of 1% lidocaine is injected for the local anesthesia, a 22-gauge, 80-mm needle will be inserted carefully in an in-plane approach from lateral to inner. Following the tip of the needle is close to the FN, a total volume 20mL of 0.375% ropivacaine will be slowly injected after negative aspiration every 5-mL. The ultrasound view of the fluid spread will be observed to ensure that the ropivacaine is injected around the FN.

### Postoperative management and follow-up

After surgery, when participants are transferred to the ward, vital signs (heart rate, noninvasive blood pressure, SpO 2 and respiratory rate) will be monitored. In 48h postoperatively, a PCIA with morphine will be begun at the end of the operation and set to the bolus only mode (bolus 1.0 mg,lockout 6 minutes,maximal does 15mg/4 hours). All participants will receive 1 g paracetamol 6 hourly as a part of postoperative multimodal analgesia. In case of nausea and vomiting, 5mg of tropisetron injection will be intravenous.

### Outcome measure

Before a series of clinical-scale evaluations and analgesia will be evaluated and recorded by the estimator, who is blinded to the group assignments, patients will be tested with the MMSE on the first and second postoperative days.

The primary outcome measurement of this study is the QoR-15 scale at 24h postoperatively. There are 15 items in the questionnaire about the quality of recovery. By being answered on an 11-point numerical rating scale, the score of each item ranges from 0 to 10. The full total scores of the QoR-15 are 150. The mean value and standard deviation will be used for descriptive analysis.

The secondary outcome are the strength of quadriceps, the VAS of the resting and the dynamic pain, the total morphine consumption, the rescue analgesic, the first time of postoperative out-of-bed mobilization, and complications. Patients will be asked to raise a straight leg of the affected limb to 15 degrees before and 3o minutes after the blockade to confirm if the quadriceps weakness is displayed. At the time before and 30 min after the block performance, 6h, 12h, 18h, 24h and 48h postoperatively, the pain scores at rest and on movement will be assessed respectively. After the patient have been resting in bed for 15 min, VAS at rest will be assessed. When VAS on movement is assessed, participants will be asked to flex hip. Being used an 11-point numerical rating scale, the VAS ranges from 0=no pain to 10=unbearable pain. The total morphine consumption in PCIA and the time of the bolus will be recorded. If the participants doesn’t feel pain relief after injection of morphine from the PCIA, the participants will receive rescue anesthetic as required by the assessment, the time and the dosage of analgesics in the first 48h will be recorded. A physiotherapist will evaluate the ability of the patients postoperatively, and record the first time patients get out of bed and do physiotherapy exercises. Postoperative complications, such as pneumonia, deep vein thrombosis, myocardial infarction, cerebrovascular accident and so on, will be recorded and treated. The complications of PENG, such as the nerve damage, the local anesthetic toxicity and the vascular puncture, will be recorded. A diagram of participant recruitment and secondary outcomes measurement is shown in Table 3.

**Table 3.**
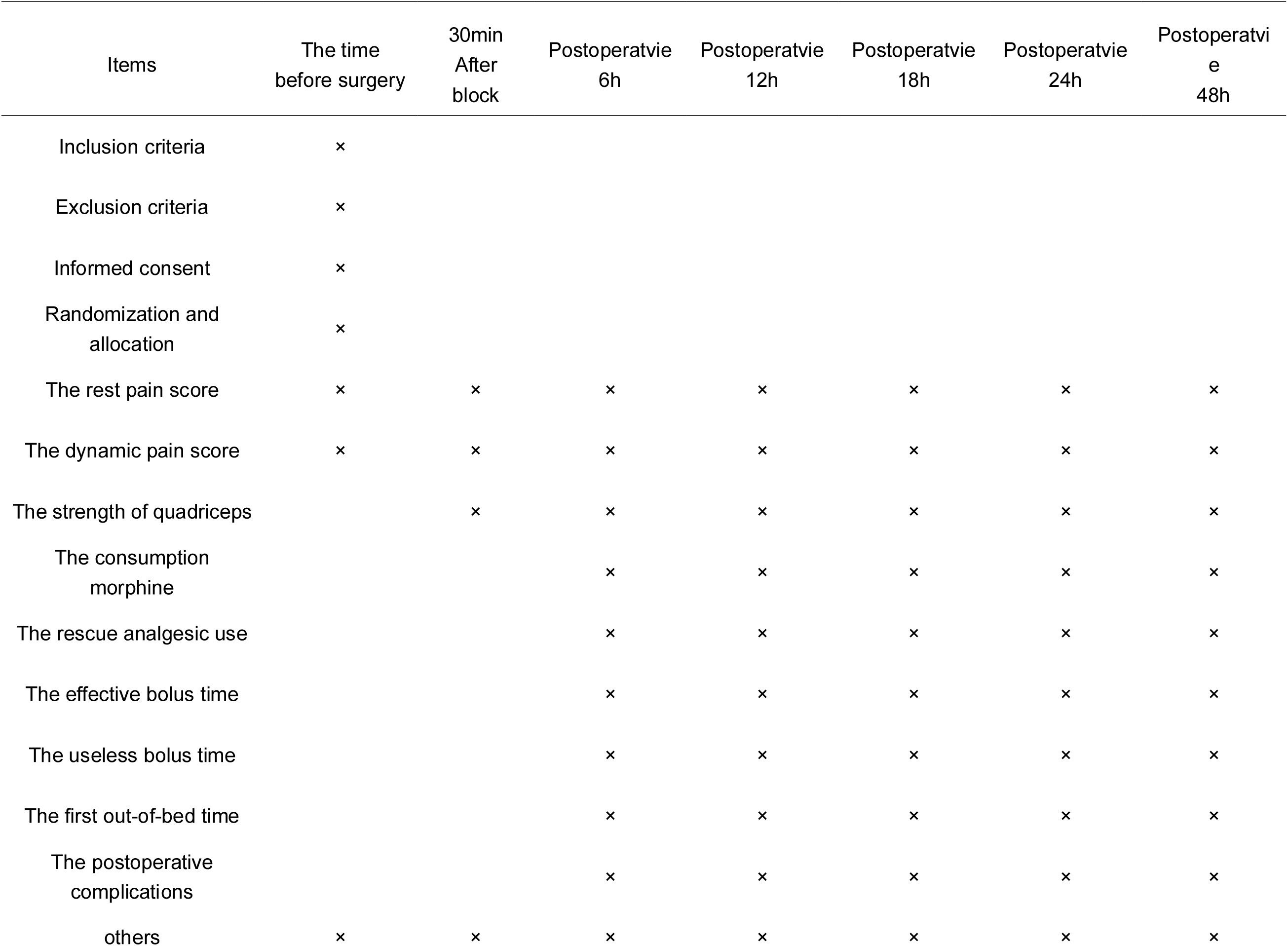
Trial process chart.

### Randomization and blinding

At the beginning of this study, a randomization sequence will be generated by SAS 9.0 statistical software and assign the participants to either the control group (the ultrasound-guided FN block group) or the intervention group(the ultrasound-guided PENG block group) on the basis of a 1:1 ratio. An independent statistician conceal the sequence (including allocation group, random numbers and intervention information) from the researcher, who assess the subjects, in the same look, opaque and sealed envelopes.

The effect of the block will be evaluated and recorded by the investigator who do not participate in the block performance. Neither the investigator nor the patients know about the grouping, and it will not be relieved until the end of the study.

### Sample size estimation

Based on the results of our preliminary experiments, the QoR-15 scores (mean difference ± SD) of elderly patients for 24h were 98.97 ± 10.37 in the PENG group and 88.25 ± 8.32 in the FN block group. Myles *et al*.^21^ found that the minimal clinically important difference for QoR-15 scale is 8.0.Therefore, to detect the effect size (power = 0.8) with the type 1 error of 5% (α=0.05), a dropout rate of 10% and a non-inferiority or superiority margin of 8, a sample size of 92 participants is required. As the result, 46 participants in each group are recruited in this study.

### Reporting of adverse events

In the period of the study, the participants will be seen daily. All adverse events will be recorded. In case of any serious complications, researchers will be informed immediately, take measure based on symptom and make discussion with any treating medical practitioners about the seriousness and reason. The sequence will be revealed. And then researchers will evaluated the relations between the serious complications and the PENG block. All details of serious adverse events will be written down and reported to the ethics committee.

### Ethics and declarations

The trial received ethical approval from The First Affiliated Hospital of Guangzhou University of Chinese Medicine Research Ethics Committee on 15 December 2020 (Reference K2020-110). This study has been registered at the Chinese Clinical Trial Registry (ChiCTR2100042341). The trial will be conducted in accordance with the Declaration of Helsinki 1996, principles of good clinical practice, and the Department of Health Research Governance Frame-work for Health and Social Care.

### Data management and monitoring

Demographic data and mental assessment data, QoR-15 data and information on pain scores, mobilization assessment data and complications will be collected and input into an electronic database. An independent researcher will guarantee the data quality during the process. All data of outcomes will be input into another independent data and double-checked to promote the data quality. The individual privacy information will be deleted to protect confidentiality. After data storage, only researchers can access to the final trial dataset directly. The progress and safety of this study will be monitored by the data monitoring committee, which composed of two professors outside the study. The DMC can give suggestions about the safety, and even pause the trial.

### Statistics analysis

All allocated subjects with available data will be analysed. According the variable type and distribution, data will be presented as mean and SD, frequency and proportion or median and IQR (25th-75th percentile). Base on the χ2 test or Fisher’s exact test, categorical variables will be evaluated. The parametric t-test, the Wilcoxon rank-sum test or the Kruskal-Wallis test will be used to analyse differences between two groups in continuous variables. And non-normal distributions will be assessed with the Mann-Whitney U test. To manage the missing data, mean completer and regression will be used. A p value of <0.05 will be considered statistically significant and results will be presented with 95% CIs. Analysis of date will be performed with the SPSS software V.21.0 (developed by IBM Corp, Armonk, New York, USA).

## Discussion

Hip fracture is often associated with serious pain. Lack of sufficient pain treatment leads to not only the deceleration of the recovery after surgery, but also the high risk of cardiovascular adverse events and long-term chronic pain. So for rehabilitation, an adequate pain treatment should be carry into effect in the perioperative period.

Opioid use could be appropriate for the requirement of pain relief after surgery. So it is still the mainstay of potent analgesia to hip fracture worldwide in the past 20 years.^22-24^ Amalie H. Simoni *et al*.^25^ pointed out about 26.8% of patients redeemed one or more opioid prescription before surgery and 61.8% received opioid therapy postoperatively. But, unfortunately, the opioid-relatied adverse events are more common to be found in elderly. It occurs in 80 percents of patients. These adverse events, including cognitive impairment, increase of fall risk and mortality, are delay rehabilitation after surgery seriously. And opioids just offer a good pain relief at rest, but are helpless in pain on movement.

Therefore, to reduce the related adverse events and improve patients’ experience, neuraxial techniques have been recommended. Consistent evidence was found to suggest that regional analgesia techniques can reduce the pain with providing the reasonable, rapid-onset and site-specific analgesia, which is more effective than traditional systemic analgesia. In addition, there is evidence that peripheral nerve block may decrease the incidence of delirium, shorten the hospital stay, reduce morbidity and mortality.^26^ Follow the development of the ultrasound guidance, the success rate of peripheral nerve block is improved.^27^ So nerve block may have an excellent effect on fast-track recovery.

Nowadays the FN block, FIB and 3-in-1 FN block are popular peripheral nerve block techniques. Unfortunately, none of these nerve block techniques are ideal to hip fracture at present. According many anatomical studies suggested, the articular branches of FN,ON and AON innervate the anterior hip joint,which plays a great role in the innervation of hip capsule.^17 28 29^ So they should be the main goal in the regional analgesia. The three main nerves can be anesthetized by 3-in-1 FN block and FIB. However the rates of the obturator block with 3-in-1 FN block are 77-80%, while the rates with FIB are 88%.^30^ Both of FIB and 3-in-1 FN block may result in failed to anesthetize ON, and the FN block also can not anesthetize ON. In addition, these three nerve block techniques may produce quadriceps weakness, which could slow mobilization and increase the incidence of falls. Therefore a new regional analgesia, which can provide a complete analgesia without significant motor dysfunction, should be put forward.

The PENG block, developed by Arango *et al*.^18^, gives a new idea of peripheral nerve block for patients with hip fracture. With performing the PENG block in 5 patients, they found Numeric Rating Scale (NRS) for rest pain in 4 cases decreased from 4 points or above to 0 point, the reduction of NRS for dynamic pain in 5cases was more than 4 points, and the median reduction of pain was 7 points. Over 100 PENG blocks have been performed by Yu *et al*. ^31^ for hip fracture and surgery, and these blocks were effective highly. Ince *et al*. ^32^ combined PENG and lumbar erector spinae plane to provide postoperative pain treatment in a 4-year-old child undergone surgery for congenital hip dyplasia, and the FLACC score in 24 hours was less than 1 point without any need for additional analgesics.

These case series and letters give the evidence about the effectiveness of PENG for hip surgery. What impressed us most is this approach provides significant dynamic pain control with a motor-sparing effect. This may give a high quality to the early mobilization as expected. So PENG seems to meet the condition for a ideal peripheral nerve block to geriatric patients with hip fracture.

To test whether PENG is effective to enhanced recovery in elderly patients with hip fracture, a variety of measurement tool should be chosen. At present, there are many clinical observational index to evaluate the effectiveness and safety of anesthesia in postoperative recovery, but most of them are focused on the physiological endpoints, such as the incidence of complications, the length of hospital stay, the mortality and so on. Though these indexes are objective and important to be measured, the evaluations from patients’ view are more humanized and also important to be assessed. So a patient-rated QoR-15 was set out by Peter *et al*. ^33^. It includes five dimensions:pain,physical comfort, physical independence, psychological support and emotional state. The score was demonstrated with the well reliability, validity, clinical acceptability, feasibility and responsiveness, and is able to differentiate between the known factors of recovery after surgery, including ambulatory surgery. Peter *et al*.^33^ suggested there was no relation between the QoR-15 and patient age, which indicates QoR-15 could be used in aged patients. And now it has been one of common used measure in orthopedic surgery. So in this study, we choose QoR-15 as the instrument of ERAS.

## Conclusion

In conclusion,to our knowledge, this is the first study using a randomized, parallel controlled, double-blind trial to compare the QoR-15 scores between ultrasound-guided FN block and ultrasound-guided PENG block, and to explore the effectiveness and safety of PENG block in elderly patients with hip fracture. Our findings may provide an new peripheral nerve analgesia for hip fracture, which could relieve pain without motor dysfunction, to accelerate recovery. This will offer clinical evidence for the optimal analgesia method in elderly patients with hip fracture.

### Strengths and limitations of this study

1. A major strength of this study is that to our knowledge, this is the first pragmatic, parallel group,randomized controlled, double-blind trial to investigate the effect of PENG block on pain control and recovery in elderly patients with hip fracture.
2. Our findings may provide an new peripheral nerve analgesia for the elderly patients with hip fracture,which could relieve pain without motor dysfunction during rehabilitation, to accelerate recovery.
3. The limitation of this study is that there isn’t any optimal biomarker about the postoperative recovery. The patient-rated QoR-15 scale could be influence by delirium, which is a common complication in elderly with hip fracture, so we perform MMSE before QoR-15 at 24h postoperatively and compare with the preoperative result of MMSE to ensure the reliability of QoR-15.
4. There is no gold standard for the pain assessment, and we choose both VAS and the morphine consumption to promote the reliability and validity of the results.

## Author contributions

WL,JTW,QHL conceived of the study, designed the study protocol and drafted the manuscript. WL and QHL wrote the manuscript. WHM is in charge of coordination and direct implementation.JHL and HYW helped to develop the study measures and analyses.WL, JHL, HYW and YHL performed the trial. YHO input the data and guarantee the data quality. HYW and YHL provided statistical knowledge for study initiation. All authors contributed to have read and approved the final manuscript.

## Patient and public involvement

Patients and/or the public were not involved in the conduct, or design, or reporting,or dissemination plans of this trial.

## Provenance and peer review

Not commissioned; externally peer reviewed.

## Additional file

SPIRIT checklist.

### The licence statement

I, the Submitting Author has the right to grant and does grant on behalf of all authors of the Work (as defined in the below author licence), an exclusive licence and/or a non-exclusive licence for contributions from authors who are: i) UK Crown employees; ii) where BMJ has agreed a CC-BY licence shall apply, and/or iii) in accordance with the terms applicable for US Federal Government officers or employees acting as part of their official duties; on a worldwide, perpetual, irrevocable, royalty-free basis to BMJ Publishing Group Ltd (“BMJ”) its licensees and where the relevant Journal is co-owned by BMJ to the co-owners of the Journal, to publish the Work in BMJ Open and any other BMJ products and to exploit all rights, as set out in our licence. The Submitting Author accepts and understands that any supply made under these terms is made by BMJ to the Submitting Author unless you are acting as an employee on behalf of your employer or a postgraduate student of an affiliated institution which is paying any applicable article publishing charge (“APC”) for Open Access articles. Where the Submitting Author wishes to make the Work available on an Open Access basis (and intends to pay the relevant APC), the terms of reuse of such Open Access shall be governed by a Creative Commons licence – details of these licences and which Creative Commons licence will apply to this Work are set out in our licence referred to above.

